# Evaluating the impact of e-registration and mHealth on institutional delivery in hazard-prone areas of Bangladesh: A protocol for a non-randomized controlled cluster trial

**DOI:** 10.1101/2022.06.30.22277099

**Authors:** Anika Tasneem Chowdhury, Sabrina Jabeen, Zeeba Zahra Sultana, Ahmed Ehsanur Rahman, Shams El Arifeen, Ahmed Hossain

## Abstract

**Background:** Despite substantial progress, Bangladesh still has a high rate of maternal deaths owing to difficulties during pregnancy, delivery, and the postpartum period. Increasing facility delivery is mandatory to reach the goal of bringing down the MMR to <70 deaths/100,000 live births by 2030. In the era of digitalization, the introduction of e-registration and mHealth may aid the government in reaching this target. The southern part of Bangladesh is a hazard-prone area, where service uptake from institutions is low. This study aimed to determine the effect of an e-registration tracking system and mHealth counseling on institutional deliveries to pregnant mothers in hazard-prone areas of southern Bangladesh.

**Methods:** We will conduct an open-label, two-arm, non-randomized controlled cluster trial for six months and use three hazard-prone areas for intervention and another three hazard-prone areas for control. We will collect data at baseline and end-line of the study period using a structured questionnaire. We will enroll at least 268 pregnant mothers from the intervention and 268 pregnant mothers from the control areas after screening based on the inclusion and exclusion criteria. Pregnancy information will be obtained from the Family Welfare Assistant register. We will follow the participants until their delivery and exclude those respondents from the study who will have post-dated delivery, migrate out, lost to follow-up, or die during the study period. Random-intercept mixed-effect logistic regression will be performed to explain the relationship of e-registration and mHealth package with institutional delivery.

**Discussion:** Institutional delivery is still uncommon in Southern Bangladesh despite several interventions. Innovative approaches like e-registration and mHealth counseling may be helpful to bring women to health facilities.

**Trial registration:** This experiment is registered in the open science framework (registration number: DOI 10.17605/OSF.IO/YZE5C) and https://www.clinicaltrials.gov/ (registration number: NCT05398978).

## Introduction

The Sustainable Development Goal-3 aimed to scale down the maternal mortality ratio to below 70 deaths per one hundred thousand live births by 2030 [1]. Despite rigorous efforts across the nations worldwide, approximately 810 women die due to obstetric complications every day [2]. In response to the call and to achieve the targeted goal of SDG, the world has strengthened the traditional approaches, adopted innovative interventions, and spread out its scope to inter-sectoral involvement. Incorporating e-registration and mHealth into the maternal healthcare system is one such step that is showing promising results in both developed and developing countries [3-4].

Institutional delivery is one of the key tools to ensure safe delivery under trained medical practitioners and tackle any delivery-related complications efficiently reducing maternal deaths and thus help to reach the SDG target. According to a joint report by World Health Organization and UNICEF, around 83% of deliveries were conducted by skilled providers worldwide during the 2014-2020 period [5]. However, the emergence of the global pandemic COVID-19 has decelerated the progress to a great extent [6]. Setting up an effective tracking system and adopting a patient-centered approach to encourage the patients for facility delivery are inevitable to confirm an increase in the institutional deliveries [4].

A non-randomized controlled study conducted by Solomon Shiferaw et al. in Ethiopia featured an increase of 14.7% in facility delivery compared to the control group due to implementing mHealth services using both voice calls and text messages [4]. Similar positive findings were observed in a study conducted in Tanzania by Julia D. Battle et al. [7].

Bangladesh targets to drop down MMR to 121 deaths per one hundred thousand live births by 2022 and below 70 deaths per one hundred thousand live births by 2030 [8-9]. Here, 53.8% of the deliveries are conducted at the facility level, whereas the target of the Government of Bangladesh is in reaching a rate of 85% by 2030 [10-11]. In the era of digitalization, the introduction of e-Registration and mHealth may aid the Government of Bangladesh to reach this ambitious target. A study conducted in Bangladesh on the impact of mHealth among Aponjon users has shown promising results. 45% of the Aponjon users went for facility delivery compared to 32% who chose non-institutional deliveries [3]. But the study interventions were e-Registration and mHealth using text messages only. There was no use of voice calls [3].

However, the effect of e-registration tracking and mHealth counseling in both voice calls and text messages as a package on institutional delivery in Bangladesh country context is yet unknown. Although the study conducted in Ethiopia showed positive results in increasing the facility delivery using both text messages and voice calls, the cultural, social, and geographical variations present between the Ethiopian and the Bangladeshi population beg the importance of a separate study to be conducted in the Bangladesh country context. Additionally, the Southern part of Bangladesh is a hazard-prone area, and the service uptake tendency from the institution is much lower here. Thus, achieving the targeted institutional delivery rate is a challenge.

In Bangladesh, paper-based registers are used in the health system to keep a record of health-related data of the care-receivers. The Directorate General of Family Planning (DGFP) of the Ministry of Health and Family Welfare (MoHFW) of Bangladesh uses Family Welfare Assistant (FWA) registers to keep track of pregnant mothers at the community level. Each FWA is assigned to one or more than one adjacent union that visits houses to conduct population registration, eligible couple registration, and pregnant mother registration quarterly. The data are stored in the paper-based registers, and union-level data are compiled monthly to produce sub-district level reports. The field activity and data validity are checked by the Family Planning Inspectors (FPI)s. However, carrying a large register during home visits, input data in the field, and monthly compilation of the huge dataset manually is cumbersome, time-consuming, and prone to human error. Digitalization of the FWA register to an e-Register system is a step forward to initiating e-registration in Bangladesh taken by the Directorate General of Family Planning in accordance with the rest of the world [9]. This initiative has eased the registration system, decreased workload, increased efficiency, and minimized human error [12].

The Management Information System (MIS) unit, DGFP started e-Registration in Chandpur Sadar, Kachua, and Faridganj sub-districts of Chandpur district with the technical assistance of Maternal and Child Health Division of icddr,b under the Safe Motherhood Promotion-Operations Research on Safe Motherhood and Newborn Survival (SMPNS) Project. The project aims to reduce maternal and newborn mortality by developing and evaluating intervention packages at the project implementation sites. Strengthening pregnancy registration is one of the key mandates of this project, and assisting DGFP in the roll-out of the eMIS registration system in Chandpur district was done to achieve the mandate. The project has employed midwives in the Chandpur District Hospital, Kachua Upazila Health Complex, and Faridganj Upazila Health Complex who will send text messages and make voice calls to counsel the registered pregnant mothers for institutional delivery. Therefore, we plan to conduct a two-arm, non-randomized controlled cluster trial to evaluate the joint effort of the DGFP eMIS tracking system and mHealth counseling done by the midwives in overcoming the challenge and improving institutional deliveries in the hazard-prone areas of Southern Bangladesh. We expect that e-Registration and mHealth intervention will provide policy-relevant lessons on scaling up of this package to improve the facility deliveries in hazard-prone areas in Southern Bangladesh.

### Objectives of the study

The primary objective of this study is to determine the effect of e-registration and mHealth on institutional deliveries in the hazard-prone areas of Southern Bangladesh. The secondary objectives are to determine the prevalence of institutional deliveries, different modes of institutional deliveries, and the birth outcomes among mothers in these areas of Bangladesh.

### Hypothesis

We hypothesize that e-registration and mHealth will increase institutional deliveries in the hazard-prone areas of Southern Bangladesh.

## Materials and methods

We used the SPIRIT checklist when writing this report and the standard protocol items are shown in **Fig 1**.

**Figure 1:**
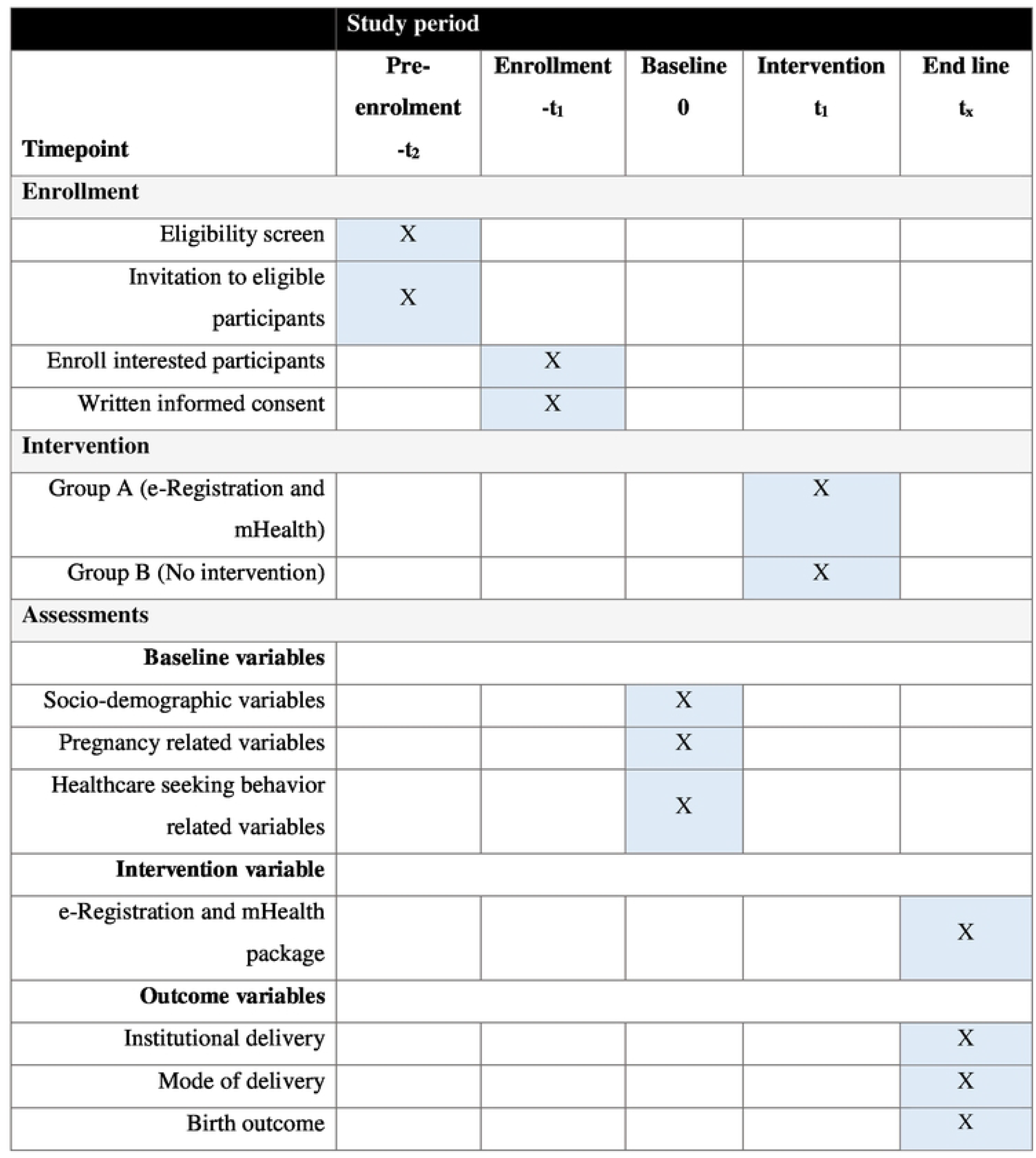
Standard Protocol Items: Recommendations for Interventional Trials (SPIRIT) checklist.

### Study design and setting

The study will be an open-label two-arm non-randomized controlled cluster trial for six months where Chandpur Sadar, Kachua, and Faridganj sub-districts will be the intervention arm as MIS unit; DGFP has initiated the eMIS registration system in these three sub-districts of Chandpur district. Chandpur is a district under the Chattogram division, located in the Southern part of Bangladesh, and is a disaster-prone area. There are 14 unions in Chandpur Sadar, 12 in Kachua, and 15 in Faridganj sub-districts of Chandpur [13]. The Government of Bangladesh has taken the initiative to implement various models in different districts, such as the Maternal, Newborn and Child Survival (MNCS) and Maternal, Newborn and Child Health (MNCH) project by UNICEF; Maternal, Newborn Health Initiative (MNHI) through joint UN initiatives; and USAID supported program MaMoni. But none has chosen Chandpur district as their project implementation site. Again, the majority of the social and health indicators of Chandpur Sadar, Faridganj, and Kachua sub-districts are below national estimates mostly due to the hazard-prone nature of these areas. So, we have chosen these three sub-districts of Chandpur district as intervention areas. The e-registration and mHealth services are not available in Bhola District. In addition, Bhola is likely to be similar to Chandpur, especially in terms of its geographical location, susceptibility to disasters, and health care and care-seeking practices of the inhabitants. So, we have selected Bhola Sadar, Charfesson, and Lalmohan sub-districts of the Bhola district as the control arm. Bhola Sadar has 13 unions, Charfesson has 14 unions, and Lalmohan has 9 unions [13]. Unions will be considered as cluster units, and individual study participants will be the sample unit.

### Registration

This experiment is also registered in the open science framework (registration number: DOI 10.17605/OSF.IO/YZE5C) and https://www.clinicaltrials.gov/ (registration number: NCT05398978, date: 26/05/2022). If any protocol modification is needed we will communicate with them. AH is the primary sponsor of this trial.

### Recruitment of the participants

The FWAs register all the pregnant mothers of their respective unions in the traditional paper-based system. We will digitalize an e-registration system in the intervention areas and in the control areas we will continue the paper-based register. The Family Planning Inspectors (FPI) then collate the data and a complete list of pregnant mothers at the sub-district level is formed.

The flow diagram of pregnant mothers’ enrolment from the FWA register is given in **Fig 2**. The list of all the existing pregnant mothers who will be in their 28 – 36 weeks of pregnancy during the first 15 days of the baseline period of both the intervention and control areas are registered in the paper-based FWA register. We will invite them to participate in the study, screen the interested participants based on our inclusion and exclusion criteria, and enroll at least 268 pregnant mothers for the intervention arm and 268 pregnant mothers for the control arm upon taking written informed consent.

**Fig 2:**
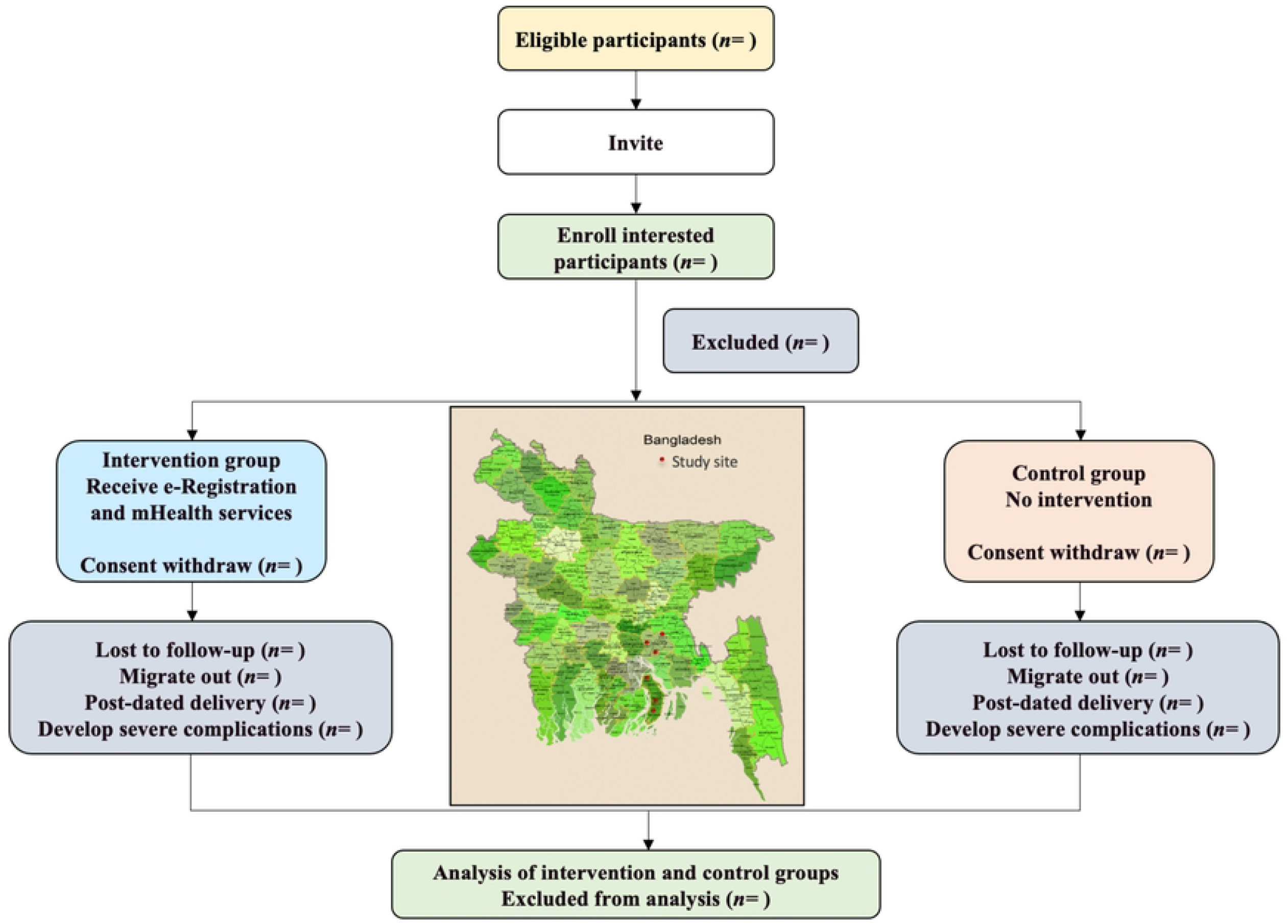
Flow diagram of pregnant mothers enrolment from FWA register.

The inclusion and exclusion criteria of the participants are given in the following:

#### Inclusion criteria

1. Pregnant mothers registered in the FWA register,
2. Age group 19 – 45 years,
3. Residence of - Intervention group: Chandpur Sadar, Faridganj, or Kachua sub-districts of Chandpur District, and Control group: Bhola Sadar, Charfesson, or Lalmohan sub-districts of Bhola District,
4. Gestational period of 28 – 36 weeks,
5. Have access to a mobile phone.

#### Exclusion criteria

1. Pregnant mothers who do not have mobile phone access,
2. Pregnant mothers with severe pregnancy-related complications at the baseline,
3. Pregnant mothers who will develop severe pregnancy-related complications, migrate out, lost to follow-up, or die during the study period,
4. Mothers who will deliver beyond 42 weeks of their gestational period,
5. Pregnant mothers who will refuse to participate in the study.

### Sample size

We have planned to recruit and then follow up a minimum of 268 pregnant mothers in the intervention areas and 268 pregnant mothers in the control areas. According to a study conducted in Ethiopia, the implementation of mHealth services using both voice calls and text messages increased facility delivery by 14.7% in the intervention group compared to the control group (43.1% compared to 28.4%; AOR: 1.98 (95%CI: 1.53, 2.55)) [4]. We decided on this proportion we need to determine the number of respondents for our study at a 5% level of error margin and 80% power. Using the formula below we determined the sample size to be 268 for each of the intervention and control groups.

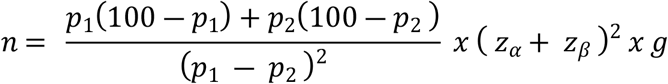

Here,

p1 = 53.8% (Proportion of institutional delivery in Bangladesh) [14]

p2 = 68.5 % (Proportion of institutional delivery following intervention assuming a 14.7% increase) [4]

z_α_ = 1.96 (at 5% error margin)

z_β_ = 0.85 (at 80% power)

g = 1.50 (design effect)

non-response rate = 5%

**n = 268**

Based on the logic, we anticipate that the sample size will have enough statistical power to meet the primary objective of our study.

### Data collection process

**Figure 3** provides the conceptual framework of data collection process. We will use a structured questionnaire developed in Bangla for data collection at the baseline and end line of the study period. Two local recruits at each of the intervention and control areas will be employed for data collection to avoid any issues with the local dialect. Among the two data collectors, one will be a female who will collect data from the pregnant mothers directly for the cultural issue and the nature of the questions asked, and another will be a male who will support the female data collector during community visits. We will train the data collectors on the questionnaire, data collection method, ethics, and infection prevention and control measures considering the COVID-19 context according to the WHO standards.

**Figure 3.**
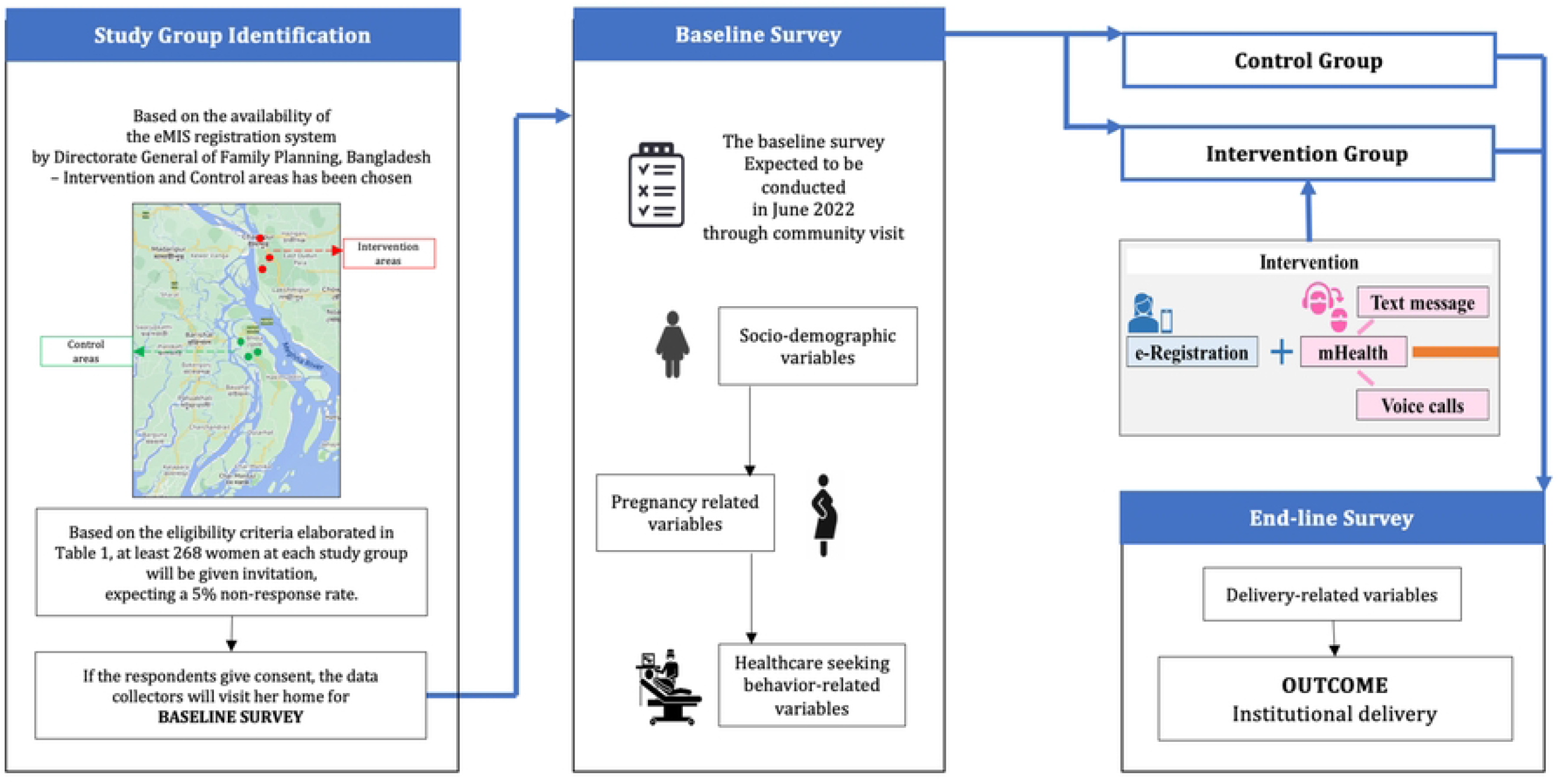
Conceptual framework of data collection process.

The questionnaire is subdivided into three main sections – screening measure, baseline survey, and end-line survey. During the baseline period, we will screen pregnant mothers using the screening tools. The screening tools will include mothers who will give consent and exclude pregnant mothers with severe pregnancy-related complications, such as eclampsia, antepartum hemorrhage, sepsis, and/or COVID-19 infection. The baseline section is designed to collect data of the respondents about the socio-demographic information, pregnancy-related information, such as parity, gravida, months of pregnancy at the first antenatal visit, counseling on facility visit, birth preparedness plan, knowledge on danger signs, location, and outcome of previous pregnancy and existing comorbidities, and other information such as personal belief, social norm, choice of delivery place, health-seeking behavior in general, and the distance between the respondent’s home, and the nearest healthcare facility. It will take approximately 20 minutes to complete the baseline section of the data collection tool. The pregnant mothers will then be followed-up until their delivery. Then their delivery-related data, such as place of delivery, mode of delivery, and the delivery outcome, will be collected at the end line of the study period using the relevant section of the questionnaire, which will take about 10 minutes to fill up.

Since the baseline, the end line data collection will be done at the end of the fourth month of the study period. The baseline and end-line data collections will be done by face-to-face interviews. The questions will be read out to the participants and the data collectors will fill up the questionnaire with the information given by the mothers. The mothers who will have post-dated delivery, develop severe pregnancy-related complications, will be lost to follow-up, or die during the study period will be excluded from the study. We will exclude post-dated deliveries and mothers with severe pregnancy-related complications as, by nature of the emergence, these deliveries are conducted in the healthcare facility and do not depend on the choice of the respondent.

### Intervention

#### e-registration and mHealth

In the intervention areas, the pregnant mothers are being registered in the FWA e-Register database so that they can be tracked and their schedules can be maintained electronically. The FWAs visit the pregnant mothers’ homes of their designated unions and register them in the eMIS database using an android tablet computer. Both online and offline data input can be done and the offline data are automatically synced to the central database once going online. The FPIs perform data quality checks, approval, and monthly aggregation of union-level data to the sub-district levels. The aggregated list will be sent to the midwives. Then the midwives will call and send text messages to the pregnant mothers whose expected delivery dates are nearby and counsel them to conduct their deliveries at the healthcare facility. The midwives will call three times in case of a failure of the first two attempts to reach each participant of the intervention arm. The text messages will be tailored beforehand in Bangla and the midwives will be trained on how to counsel the pregnant mothers on facility delivery. Following e-Registration, the mHealth intervention will be given once to the pregnant mothers of the intervention arm during 37-40 weeks of their pregnancy. We expect to evaluate the combined effect of e-Registration tracking and mHealth counseling through this study.

### Outcome measures

#### Primary outcome

##### 1. Institutional delivery

Normal vaginal deliveries, cesarean sections and assisted vaginal deliveries (forceps and vacuum deliveries) conducted at any healthcare facility by a medically trained provider/ skilled birth attendant situated at the study sites will be considered as institutional delivery.

Indicator: Proportion of institutional delivery

Denominator: Total deliveries (A sum of normal vaginal deliveries, cesarean sections and assisted vaginal deliveries)

#### Secondary outcome

##### 1. Mode of delivery

Indicator: Proportion of normal vaginal deliveries Proportion of cesarean sections

Proportion of forceps and vacuum deliveries Denominator: Total deliveries

##### 2. Birth outcome

Indicator: Proportion of live births

Proportion of stillbirths Denominator: Total births

### Data management and analysis plan

We will analyze the data in R 4.1.2. We will do a frequency distribution and proportion analysis for categorical variables, and for continuous variables, we will perform a mean and standard deviation analysis. Finally, a random-intercept mixed-effect logistic regression model will be fitted to explain the relationship of e-Registration and mHealth package with institutional delivery, where the union will be included as the random intercept. An adjusted odds ratio with p-value less than 0.05 will be considered statistically significant for the variable.

### Quality control and quality assurance

The data monitoring committee will regularly check data and monitor to maintain the data quality. An expert will double-check the validation of data collected by the data collectors. The data entry team will link records that have been collected with the help of individual identifiers assigned. We will remove this identifying information once the records are done matching. The anonymized data will be uploaded to a central web-based platform regularly, after following all the appropriate confidentiality requirements. We will use an encrypted, password-protected centralized database system to store data and only authorized members of the research team will be allowed access.

### Patient and public involvement statement

There were no funds or time allocated for the patient and public involvement, so we will not be able to involve patients. We will ask pregnant women to assist us in evaluating an effective digital health strategy to reduce adverse events.

## Discussion

This two-arm non-randomized controlled cluster trial of six months will determine the effect of e-registration tracking and mHealth counseling as a complete package on institutional deliveries in the hazard-prone areas of Southern Bangladesh. A minimum of 268 pregnant mothers of 28-36 weeks will be enrolled and followed up until their deliveries in each intervention and control area. Baseline and end-line data will be used to synthesize the study result. Since the end-line data will be collected four months later than baseline data collection, the study might have a recall bias.

However, to our best knowledge, this two-arm non-randomized controlled cluster trial will be the first study that will evaluate the effect of e-Registration and mHealth as a complete package on institutional deliveries in the hazard-prone areas of Southern Bangladesh. e-registration and mHealth intervention package may increase facility delivery, thus, reduce delivery-related complications and maternal mortality. Thus, our study will facilitate the evidence gap related to the effectiveness of e-registration and mHealth on facility delivery and provide substantial evidence which may help the government to take the necessary steps to roll out e-registration and mHealth counseling as a complete package to increase institutional deliveries and reduce the burden of maternal mortality ratio of Bangladesh by 2030.

### Trial Status

The protocol version is 1.0 (3 April 2022). We expect to recruit the participants by the first half of June 2022 and complete the intervention by the end of August 2022. End line survey will be completed by November 2022 and the result will be available in December 2022. We plan to publish the study findings in a peer-reviewed journal.

## Data Availability

No datasets were generated or analysed during the current study. All relevant data from this study will be made available upon study completion.

## List of abbreviations

ANC: Antenatal care
COVID-19: Coronavirus Disease-2019
DGFP: Directorate General of Family Planning
FWA: Family Welfare Assistant
MDG: Millennium Development Goal
MMR: Maternal Mortality Ratio
SDG: Sustainable Development Goal

## Declarations

### Ethics approval

Ethical approval has been taken from the Ethical Review Board of North South University (Ref: 2022/0R-NSU/IRB/0205) (S1 File). If any protocol modification is needed we will communicate with them. After explaining the objectives of the study, the rights of the respondent, and ensuring privacy and confidentiality, we will take written informed consent from the study respondents before data collection. The respondents will have the full right to leave the study at any time during the study period. The data collectors will maintain all necessary precautions (use mask, hand sanitizers and maintain social distance) during data collection.

### Patient consent for publication

Not required.

### Availability of data and materials

Data sharing is not applicable to this article as no datasets were generated or analysed during the current study. However, we will share make the data available upon completion of the study.

### Competing interests

The authors do not have any financial or any other competing interests.

### Funding

The work is funded by Japan’s Debt Relief Grant Aid (DRGA) Counterpart Fund (Grant# 223004100). The funder have no role in the research.

### Authors’ contributions

ATC and AH drafted the manuscript. All the authors participated in the study design and critical review of the manuscript.

## Acknowledgements

Not required.

## Supporting information

**S1 file. Ethical approval**

**S2 file: Complete protocol submitted to the ethical approval board**

**S3 file: SPIRIT checklist**

## Notes

### Competing Interest Statement

The authors have declared that no competing interests exist.

### Clinical Trial

NCT05398978

### Clinical Protocols

https://www.clinicaltrials.gov/ct2/show/NCT05398978?term=eregistration&draw=2&rank=1

### Funding Statement

SEA received the Japan's Debt Relief Grant Aid (DRGA) Counterpart Fund (Grant# 223004100). https://www.jica.go.jp/english/index.html The funder had and will not have a role in study design, data collection and analysis, decision to publish, or preparation of the manuscript.

### Author Declarations

Ethical approval has been taken from the Ethical Review Board of North South University (Ref: 2022/0R-NSU/IRB/0205)

